# Retarded Logistic Equation as a Universal Dynamic Model for the Spread of COVID-19

**DOI:** 10.1101/2020.06.09.20126573

**Authors:** B Shayak, Mohit M Sharma

## Abstract

In this work we propose the retarded logistic equation as a dynamic model for the spread of COVID-19 all over the world. This equation accounts for asymptomatic transmission, pre-symptomatic or latent transmission as well as contact tracing and isolation, and leads to a transparent definition of the instantaneous reproduction number *R*. For different parameter values, the model equation admits different classes of solutions. These solution classes correspond to, inter alia, containment of the outbreak via public health measures, exponential growth despite public health measures, containment despite reopening and second wave following reopening. We believe that the spread of COVID in every localized area such as a city, district or county can be accounted for by one of our solution classes. In regions where *R* > 1 initially despite aggressive epidemic management efforts, we find that if the mitigation measures are sustained, then it is still possible for *R* to dip below unity when far less than the region’s entire population is affected, and from that point onwards the outbreak can be driven to extinction in time. We call this phenomenon partial herd immunity. Our analysis indicates that COVID-19 is an extremely vicious and unpredictable disease which poses unique challenges for public health authorities, on account of which “case races” among various countries and states do not serve any purpose and present delusive appearances while ignoring significant determinants.

---

We have already given detailed descriptions of the origins etc of the Coronavirus in our previous two papers [1], [2] and shall dispense with such material now. We shall only mention that as we write this, there are more than 6.1 million corona cases worldwide and more than 3,70,000 deaths. Due to the considerable length of this Article, we give here an outline of its contents. We have divided the Article into three main Parts and in each Part we have several Sections. In Part 1, we first mention a paradox we have observed in the spread of the virus over the past few weeks (§1) and then present a qualitative resolution of this paradox (§2). In Part 2 we present the mathematical model proposed in this Article. Starting with a discussion of our old model (§3), we then derive the retarded logistic equation (§4). Having proposed the equation, we first validate it through data fits (§5) and then describe in considerable detail the various kinds of solutions which it can support for different parameter values (§6). This is followed by a demonstration of the distortive effects of limited testing capacity (§7) and a discussion of how all these effects combine to explain the corona spreading dynamics on the global scale (§8). From the technical viewpoint, §3, §4 and §6 form the heart of this Article and are the Sections of greatest interest to the mathematical modeler. In Part 3, we expand on the consequences of our model. Our first consideration is reopening (§9), and then we go on to effective contact tracing (§10) and ensuring public compliance (§11) which are both prerequisites. We then present an observation related to racial anomaly in the spread of the virus (§12), discuss the implications of the mathematical results on people’s behaviour (§13) and finally wrap things up with a brief conclusion. To the extent possible, we have tried to ensure that the various Sections are independent of each other, though almost the entire discussion is contingent on the model and results of §3, §4 and §6.

## 1 A PARADOX AND ITS RESOLUTION

This Part does what the title says – it describes the paradox and its solution, in mostly qualitative terms. The maths will come in the next Part.

### §1 PARADOX

When we wrote Reference [2] six weeks back, we were hopeful that lockdowns and attendant public health measures such as sanitization and contact tracing would be successful at beating corona. With as much fanfare as we could arrange for, we proclaimed our discovery of a new route to the end of COVID-19, which we called self-burnout. We presented a delay differential equation (DDE) for the case dynamics and found that it admits stable solutions for parameter values which correspond to a lockdown, even a partial one. We predicted that (insofar as such prediction is possible from a lumped parameter model) South Korea would eradicate the virus in approximately the first week of May, and that Austria would soon follow suit. We also expressed the hope that trouble spots in lockdown such as the cities of New Delhi and Mumbai in India would pretty soon bend over their curves and clamber out of the epidemic.

Over the intervening period, some of these predictions came true while others turned out to be dramatically false. South Korea did indeed achieve zero domestic cases for three consecutive days beginning on 04 May. After that it prematurely restarted normal life in the country, which led to a second outbreak as was to be expected; this renewed outbreak has again been controlled. Austria also slowed the rates further until a lockdown relaxation in late May had a dampening effect, which was again not surprising. Delhi and Mumbai however presented staggering shocks – even as residents remained under house arrest, the case counts in both cities skyrocketed [3]. Nor was India the only country to display such anomalous behaviour. Russia, which in mid-April was barely on the radar and already in a state-enforced lockdown, went up from thousands to lakhs [4] in just one month. Canada had entered lockdown in mid to late March and quickly flattened the exponential growth to linear. But it remained that way for all of the next two months, steadily adding 1000-1500 cases every day until last week, when the total was 85,000. Only now are there some signs of the curve bending back. In USA, the state of New York had declared a lockdown on 22 March at 15,000 cases, after which the counts ballooned before coming down. The explosion in cases was attributed to delay on the part of the authorities in adopting social restrictions. This apparent laxity was not present in California where lockdown had been imposed one week earlier at only 600 cases. While that state did not burst like a bomb, it embarked on a not-so-slow and very steady linear increase of cases, reaching 1,15,000 today with no end in sight. A linear but equally persistent case growth has also been manifested in Sweden which, unlike the territories mentioned before, has consciously decided to remain open and make a bid for herd immunity. These observations seemed almost beyond explanation.

Looking through the data sets, we found a second major contradiction in the plots of case rate vis-à-vis death rate in hotspot regions. It is well known that COVID-19 is a disease with a long time to death – typically the patient worsens after a week, gets admitted to hospital, deteriorates, gets put on ventilator and eventually gives up an unequal struggle. A recent publication [5] (due to topicality of the issue, almost all our technical References will be preprints; we have not tracked if some of them might have been published in the interim) estimates the time to death as approximately 20 days. Hence, in hard hit regions where cases are so numerous as to smooth out stochastic effects, we would expect the peak in deaths to come approximately 20 days after the peak in cases. In many regions however, this is not the situation. Below we show the rates of cases/day and deaths/day as a function of time for New York State, Turkey, Italy and Germany. In each plot, a green bar denotes the additional case count and a red bar denotes the additional death count for that day. All plots run upto 27 May 2020. Germany and Italy start from 20 February 2020, New York from 01 March 2020 and Turkey from 11 March 2020. In each plot, we have smoothed out the original data by taking a three-day running average twice. Finally, we have scaled up the death rate by a numerical factor so that the death bars acquire approximately the same height as the case bars – this factor is 12 for New York, 7 for Italy, 33 for Turkey and 20 for Germany.

In New York, the deaths lag the cases by about a week during the rising phase and peak about 4 to 5 days after the cases. During the falling phase, the two are almost parallel. In Turkey also, the interval between the green and red peaks is about the same although the lag is a little harder to discern. In Italy, the delay between the two is very well-defined, and again it is barely 3-4 days – indeed, the case and death counts appear to go almost hand in hand. This discrepancy has also been noted by del Re and Meridiani [6]. Only in Germany can we see a clear 15-day gap or more between the rise, the peak as well as the fall of these two quantities. While excessively rapid death may still be explainable in a region with non-existent healthcare facilities, none of the territories in Fig. 1 suffer from this problem. Therefore, something else is clearly afoot.

**Figure 1:**
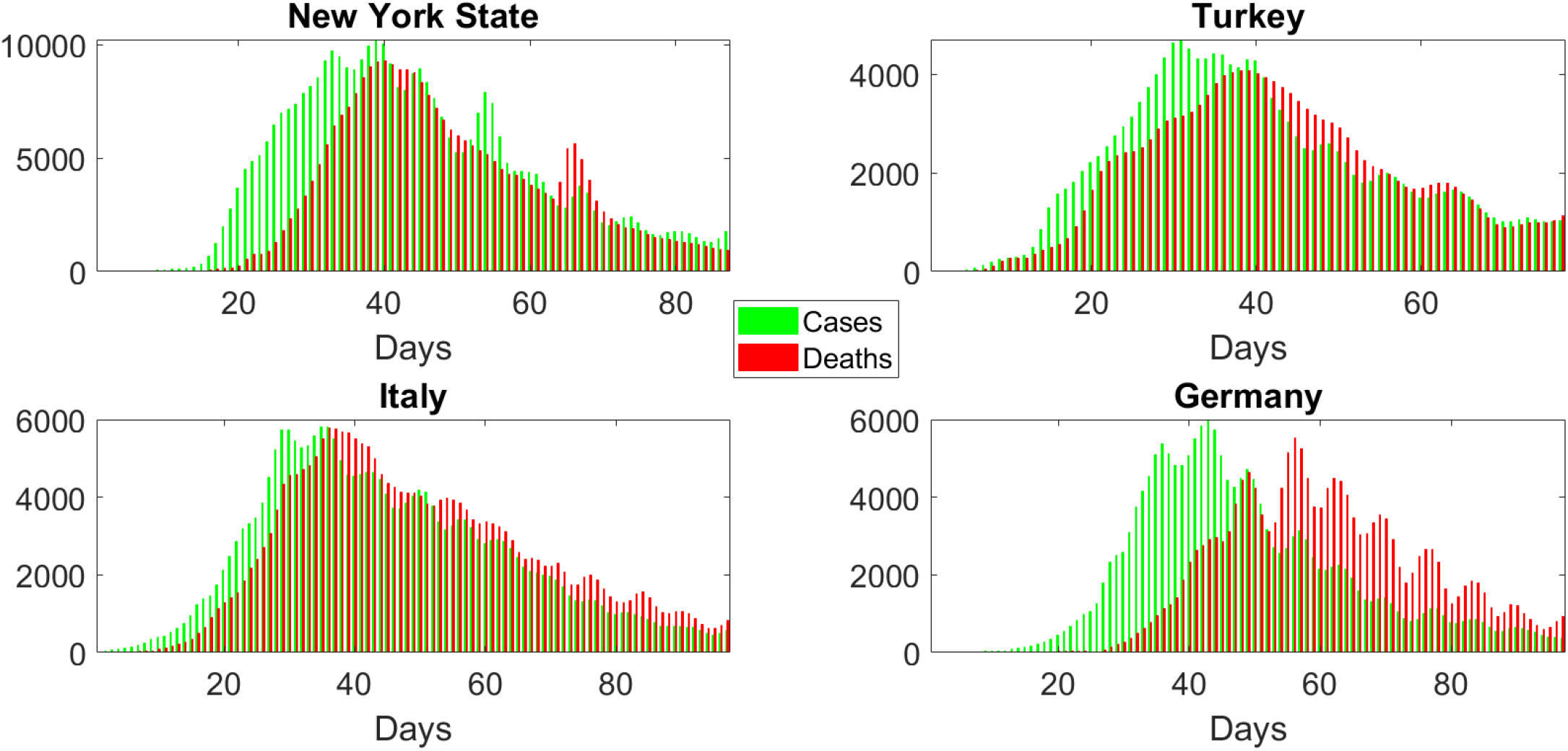
Case rate (green bars) and death rate (red bars, scaled up for visual clarity) in four regions of the world as functions of time.

**Figure 2:**
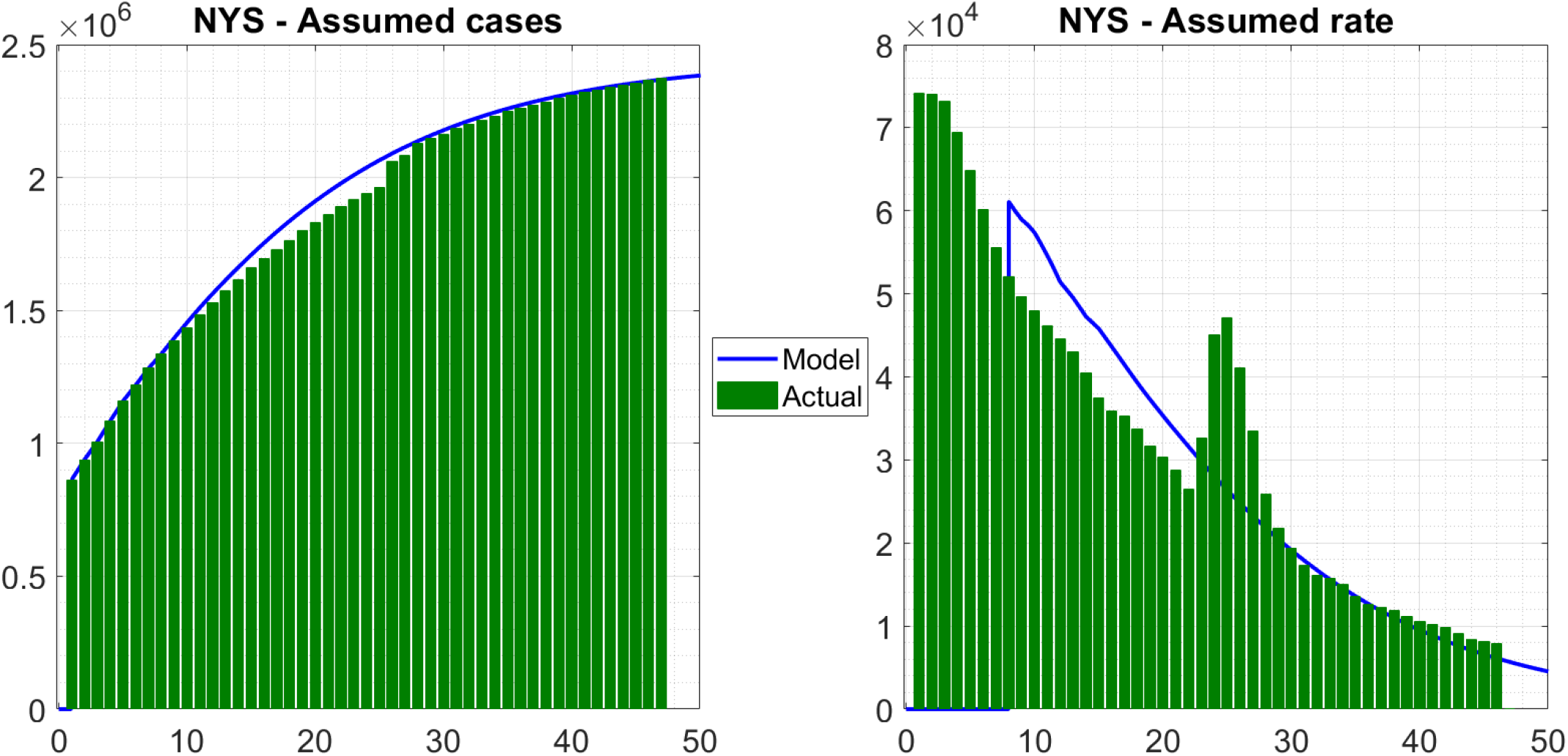
Case count and case rate predictions for New York State obtained from (3) compared with assumed actual case histories. Note that the assumed actual counts are derived by extrapolation from the reported death data.

**Figure 3:**
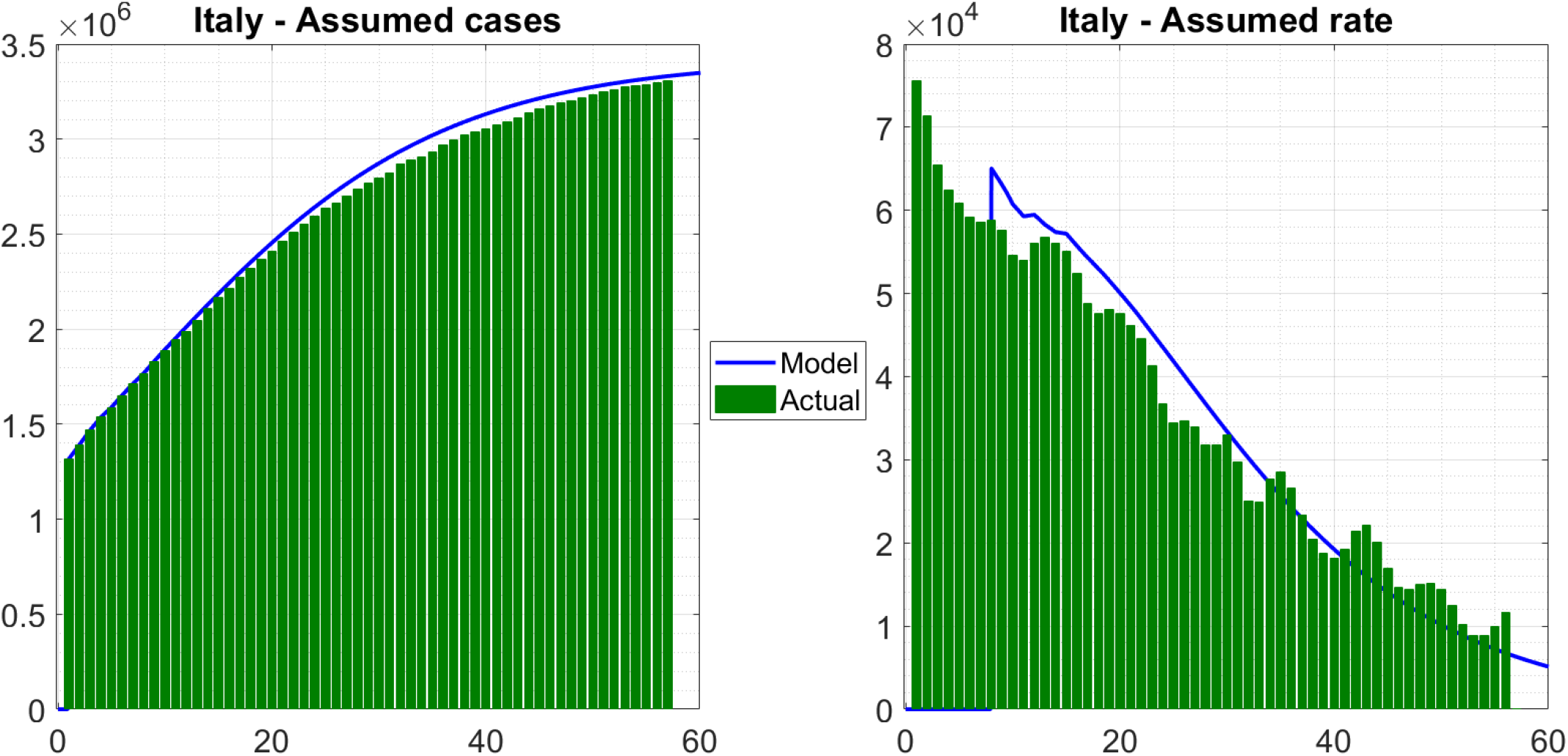
Case count and case rate predictions for Italy obtained from (3) compared with assumed actual case histories. Note that the assumed actual counts are derived by extrapolation from the reported death data.

**Figure 4:**
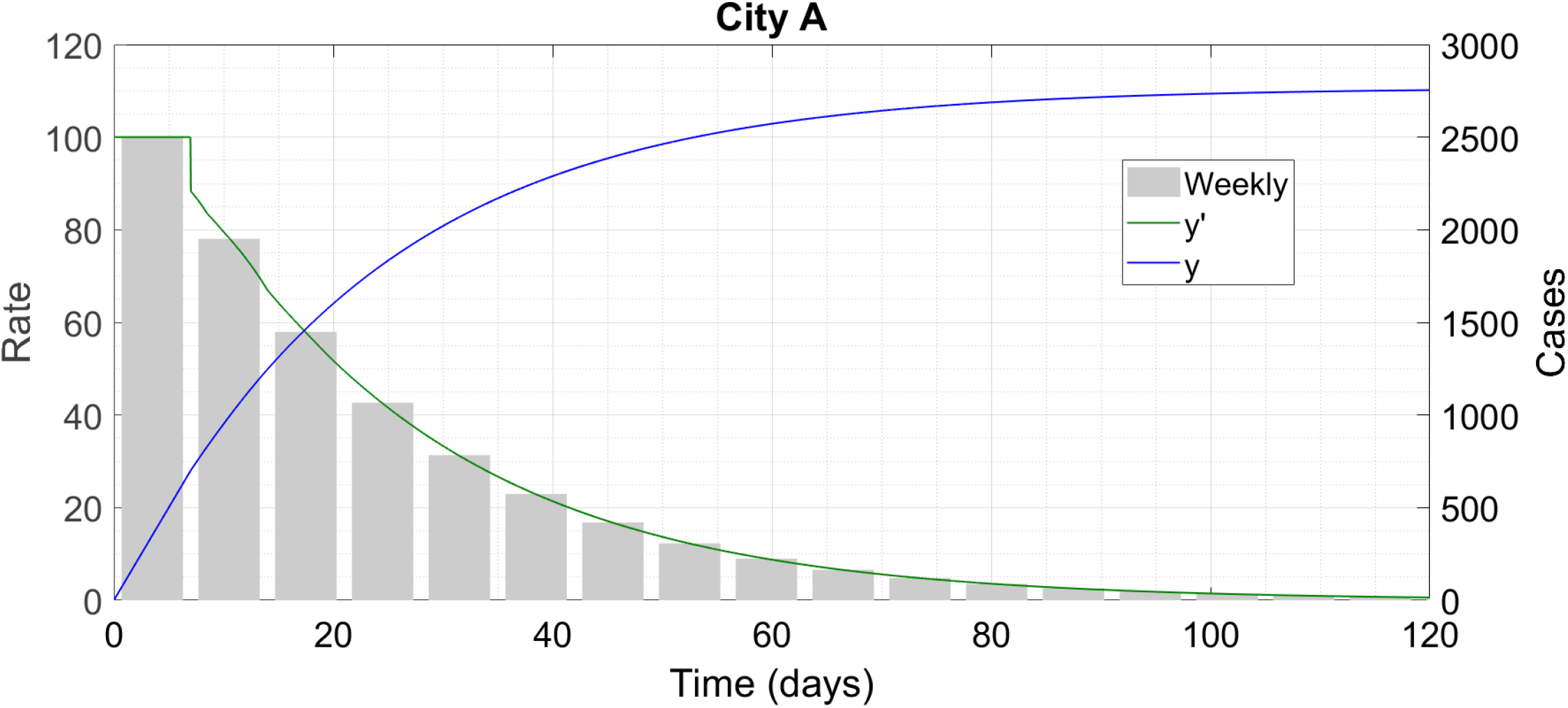
City A proceeds to self-burnout right from the start.

**Figure 5:**
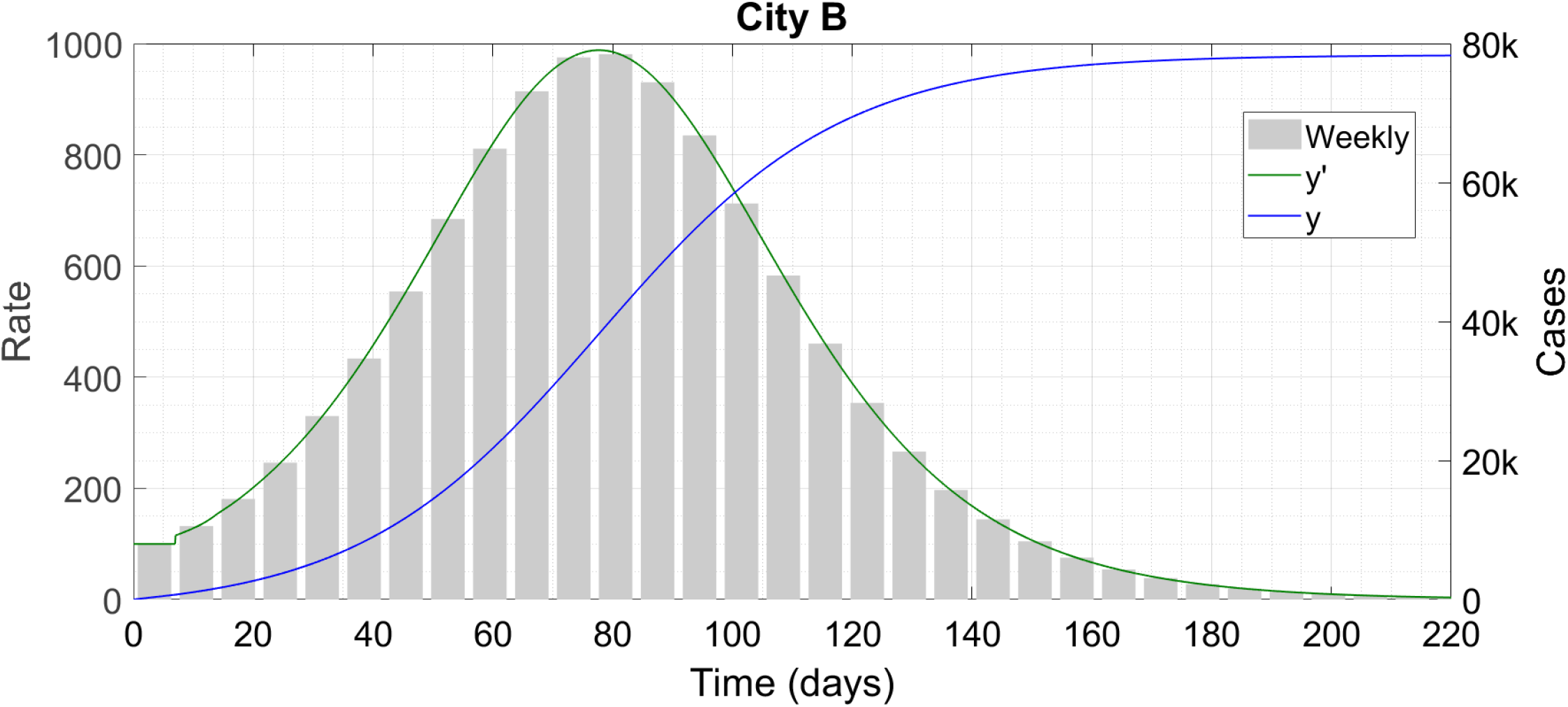
City B grows at first before reaching burnout. Note that ‘k’ denotes thousand.

**Figure 6:**
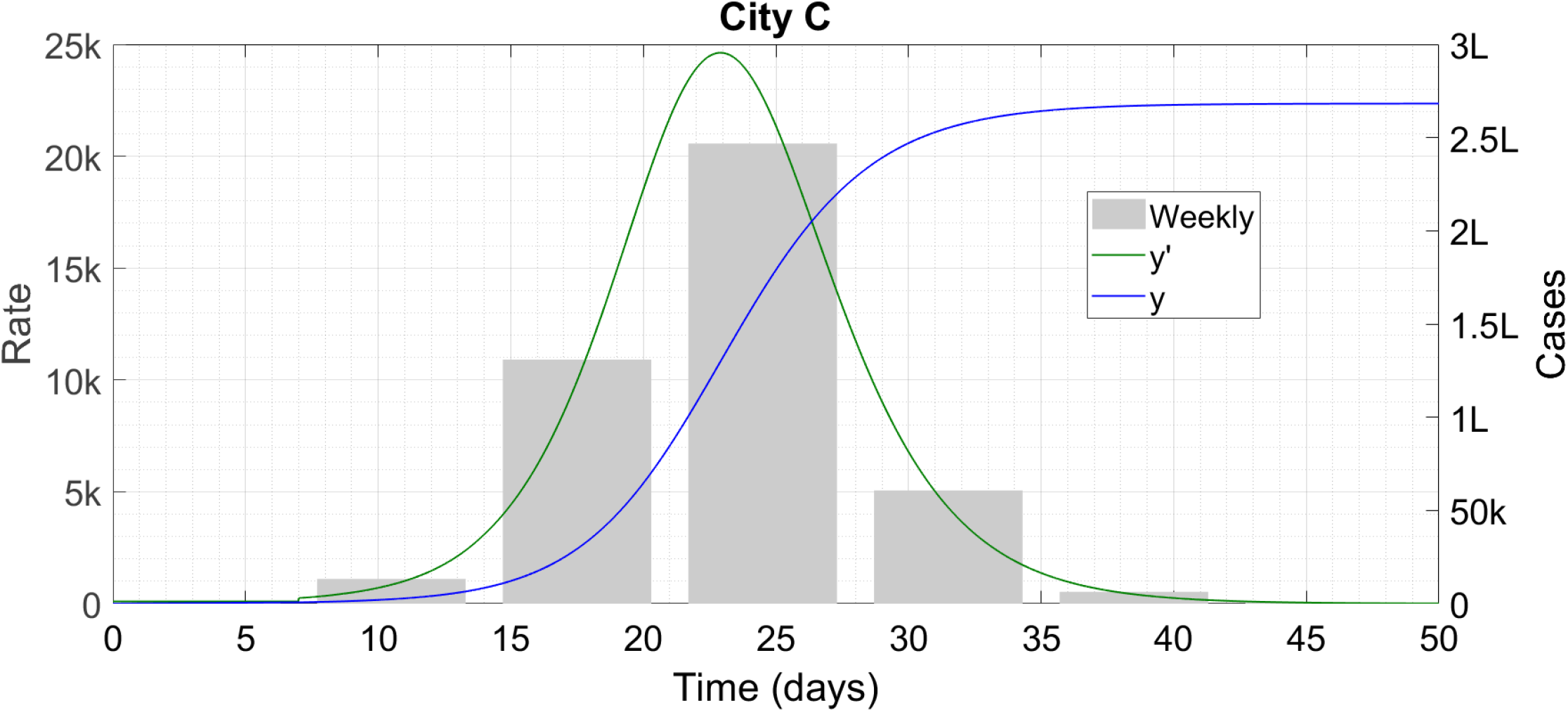
City C proceeds all the way to herd immunity. Note that ‘k’ denotes thousand and ‘L’ lakh.

**Figure 7:**
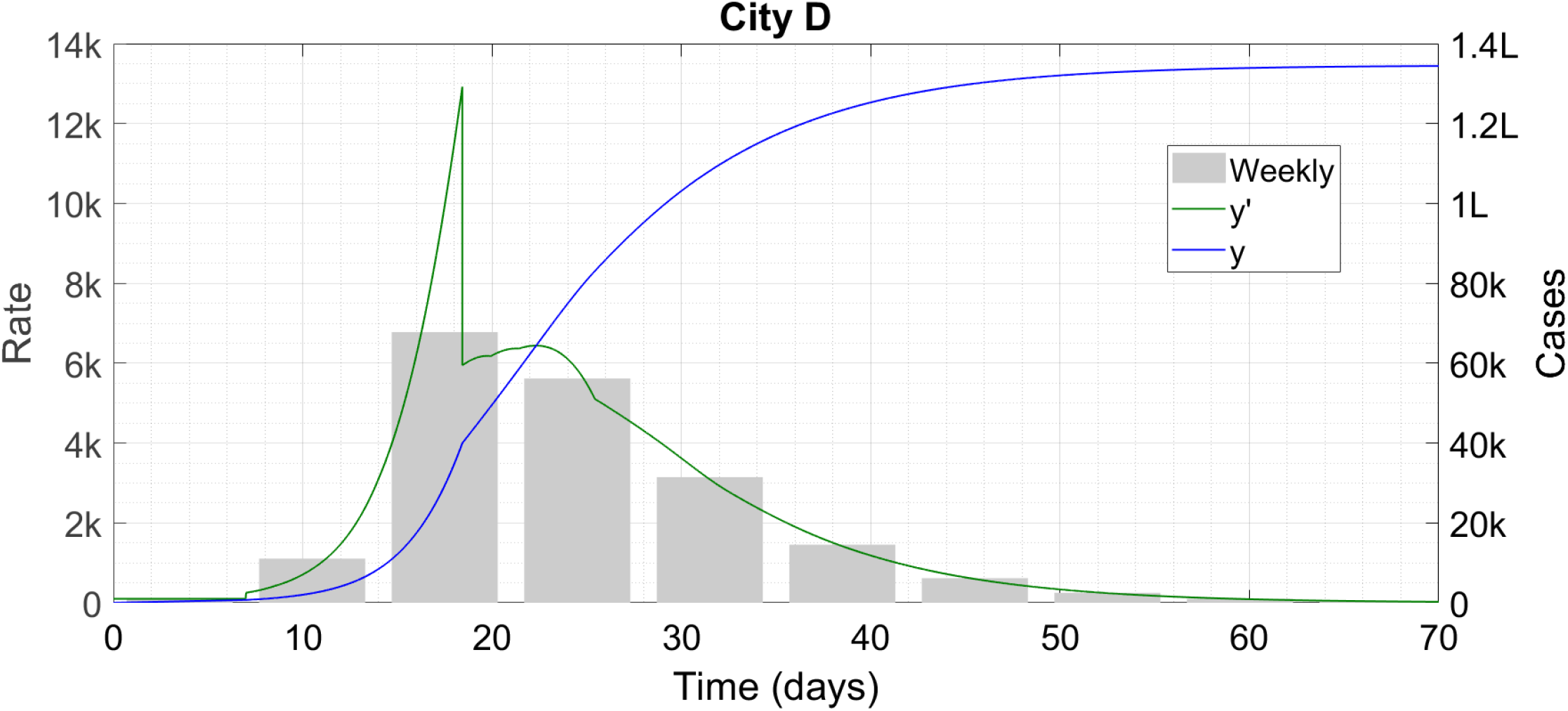
City D is a mixture of B and C. Note that ‘k’ denotes thousand and ‘L’ lakh.

### §2 RESOLUTION

We claim that asymptomatic carriers are at the root of the above conflicts. Since the infection period for an asymptomatic carrier is typically 7 days, such a person can spread the disease to a large number of people even if his/her mobility is very restricted on an absolute scale. Indeed, in the DDE presented in Ref. [2], which we are going to use in the present Article as well with a slight modification, we find that a huge contribution to the reproduction number *R* comes from the asymptomatic carriers. This happens because an undetected asymptomatic carrier can spread the disease for as long as a week, whereas the latent infectious period for a symptomatic carrier is 3 days or less. The greater the fraction of asymptomatics, the more strict are the social mobility restrictions needed to ensure *R* < 1; above a certain fraction, the required mobility level becomes impossible to achieve in real society. This can explain the growth of the disease against a lockdown. The only term which can suppress the asymptomatic effect is the contact tracing term – if contact tracing is so effective as to identify and quarantine a sizeable fraction of these carriers before they see too much of society then *R* can still be kept to a sub-unity level. This is probably what is happening in regions where the epidemic is well under control.

In regions where death tolls peak only a few days after case counts, we claim that asymptomatic carriers work in conjunction with initially low testing rates to produce this effect. In New York State for example, we contend that the actual peak of the case histories still occurred about 20 days before the death peak, but that peak remained invisible since many of the cases were asymptomatic and testing capacity was not nearly enough to even consider people with no or mild symptoms. This problem did not occur in Germany since the government there had built up huge testing capacity very quickly. This enabled the Germans to detect far more cases in the early phases than the hotspot regions which quickly got overwhelmed.

Indeed, the fraction of asymptomatic carriers being reported has gone up steadily over the past couple of months. Mizumoto et. al. [7] which we used as benchmark in our prior works [1], [2] had quoted 18 percent from an observational cohort study. This went up to 40-plus percent in a study which appeared in late April. It has still further escalated to 70 percent or more in reports issuing from Mumbai [8], [9] and Delhi [10]. In the technical literature, Thorne [11] has used asymptomatic carriers to explain the anomalous mortality rates which we have mentioned for New York and Italy, although the analysis is somewhat simplified. Kurita et. al. [12] again use asymptomatic carriers to explain anomalies in the corona dynamics in Japan and in Wuhan, China. Using a mathematical model, they estimate the fraction of asymptomatic carriers to be greater than 99 percent, which is probably somewhat excessive. Both these works find that the disease is ending through herd immunity. A very recent study by Pal and Bhattacharjee [13] also finds that asymptomatic carriers can significantly outnumber the symptomatic ones under suitable circumstances.

We now delve into the issue of the spread of corona in a quantitative manner.

## 2 MATHEMATICAL MODEL AND SOLUTION

Our starting point is the self-burnout model from Ref. [2], which we now recapitulate briefly.

### §3 THE ORIGINAL MATHEMATICAL MODEL

The DDE derived in Ref. [2] is

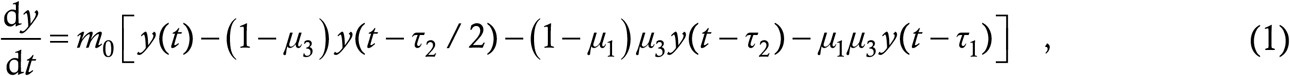

where the variables and parameters are as follows :

- *y* (*t*) is the cumulative number of corona cases in a region
- *m*_0_ is the rate (persons per day) at which every infected person spreads the infection to other people
- *τ*_1_, which we take as 7 days throughout, is the duration for which an asymptomatic carrier remains transmissible
- *τ*_2_, which we take as 3 days throughout, is the latency or pre-symptomaticity period i.e. the duration for which a person is transmissible before developing symptoms
- *μ*_1_, a number between 0 and 1, is the fraction of total cases who are asymptomatic
- *μ*_3_, a number between 0 and 1, is the fraction of total cases who fail to get detected by contact tracing and hence escape from the resulting mandatory quarantine

The values of *τ*_1_ and *τ*_2_ are in agreement with Ref. [5]. Briefly, (1) assumes that every at-large case transmits the disease to others at a constant rate *m*_0_. Symptomatic cases enter quarantine as soon as they develop symptoms, undetected asymptomatic cases transmit the disease for *τ*_1_ days while undetected pre-symptomatic or latent cases transmit for *τ*_2_ days. The model assumes forward contact tracing starting from symptomatic cases i.e. the contacts of every new symptomatic case, and possibly the contacts of these contacts etc are rounded up, tested and quarantined but no attempt is made to determine wherefrom the original case him/her-self contracted the infection. This testing and contact tracing activity is treated as instantaneously capturing a fraction 1 − *μ*_3_ of all the symptomatic as well as asymptomatic cases. For more details of the derivation, please consult Ref. [2].

That reference also obtains the criterion for stability of the solutions of the above equation. Indeed, the reproduction number *R* comes out naturally from (1) as

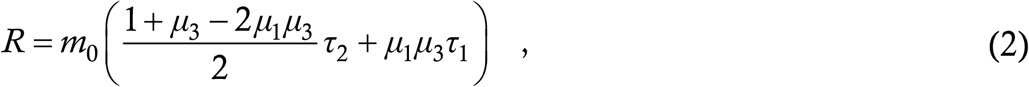

with the epidemic terminating in time if *R* < 1, growing exponentially if *R* >1 and growing linearly if *R* = 1 (for the details, again consult Ref. [2]). In Ref. [2] we had used the value *μ*_1_ = 1/5 i.e. 20 percent of cases are asymptomatic, as was current knowledge at that time. Then, assuming *μ*_3_ = 1/2 i.e. 50 percent effective contact tracing, which seems plausible (see also the next paragraph), we had found a critical *m*_0_ of 20/53 or 0.3774. In a lockdown where the average person steps out of his/her house only once in 4 to 5 days or so for necessities, this criterion had ample margin to account for high-interaction essential personnel such as grocers and food workers. As such, it did not appear particularly hard to meet, motivating our conclusion that the epidemic could be driven to extinction everywhere by mobility restrictions, six-feet separation minima (an aeronautical term which we personally prefer to “social distancing” since it does not have any connotations of emotional isolation), sanitization and preventive testing. If *μ*_1_ is raised from 0.2 to 0.8 however, the critical *m*_0_ drops to 0.2597, which is dangerously close to the minimum possible rate at which one has to go out. Adding the contribution of high-interaction essential workers and the absence of preventive testing in most societies, we are very likely staring at exponential growth of the outbreak even under the most rigidly enforced lockdown.

The only effect which can still prevent the spread of the disease is contact tracing. For in (2) we can see that the asymptomatic contribution to *R* is *μ*_1_*μ*_3_*τ*_1_. Even if *μ*_1_ is high, a sufficiently low *μ*_3_ can still keep the size of this term small and result in an *m*_0_ which is practical to achieve in real life. For example, the *m*_0_ of 0.37 which we had used in Ref. [2] can be obtained with *μ*_1_ = 0.8 if *μ*_3_ is lowered to about 0.25 i.e. if 75 percent of all infected people, including asymptomatic carriers, can be captured through contact tracing. Thus, it is likely that countries such as Germany, Austria and Switzerland, which have fared considerably better than otherwise similar countries such as France, Belgium and the UK, have done an excellent job of contact tracing. The pivotal importance of contact tracing has also been highlighted by Cherednik [14], [15] who (using a logic different from ours) considers quarantine as “hard measures” and separation, sanitization etc. as “soft measures” in the control of the epidemic. Very recently, we have obtained the first estimates of *μ*_3_from an official source. The Tompkins County Health Department has released figures of the spread of COVID in Tompkins County, New York State [16]. Their figures indicate that 86 of the county’s 154 cases could be traced as having had exposure to a known case. This gives a value of *μ*_3_ slightly less than 1/2. However, it is very likely true that all the cases which did not emerge from contact tracing were symptomatic, and there must be many asymptomatic cases also which have not been detected at all in contact tracing drives. Thus, the actual *μ*_3_for Tompkins County is in all probability greater than 1/2.

Equation (1) suffers from one obvious limitation. This is that, since it is a linear equation, if *R* > 1 then the disease keeps growing exponentially. This growth however cannot continue indefinitely – at some point *y* will become equal to the region’s entire population, at which point (if not long before) the growth must caese completely. In Ref. [2] we did not account for this fact since we were interested only in the regime where *R* < 1. Now however, we have to add a correctional term which allows for a departure from exponential growth after the case load becomes sufficiently high.

### §4 THE NEW PROPOSED MATHEMATICAL MODEL

The correction to (1) comes from the *m*_0_ term. As already mentioned, this term means that each at-large case spreads the disease to *m*_0_ people every day. As discussed in Ref. [2], two factors contribute to *m*_0_ – the rate at which the case interacts with other people and the probability that each such interaction actually results in transmission. For example, if an undetected case goes out once every 3 days, interacts with one person on each trip, and each interaction carries a 50 percent transmission probability, then the contribution of that case to *m*_0_ is 1/6. We assume that each person can only be infected once i.e. a recovered case cannot be reinfected by Coronavirus. Although there have been occasional reports of such reinfection, the credibility of those is not established beyond doubt [17], [18] and the overall reinfection probability still appears negligibly small. A very recent reference [19] finds that immunity against benign coronaviruses (NOT the current, dangerous strain) lasts for about one year, which is enough time for the epidemic to be brought under control. Accordingly we shall stick to the assumption of single infection in this Article.

Now, if the social mobility remains constant, then what happens as the epidemic evolves in a region is that the interaction rate of each new case with other people remains the same, but a greater and greater fraction of these people happens to be recovered cases who cannot be infected any longer. Thus, over and above the interaction rate and the basic transmission probability, a third probabilistic contribution adds on to *m*_0_ – the probability that the person with whom the case is interacting is not already a recovered case. This probability is 1 − *y*/*N* where *N* is the total susceptible population of the region. Accordingly, as a first correction, *m*_0_ in (1) may be replaced by *m*_0_ (1 − *y*/*N*) to give the revised DDE

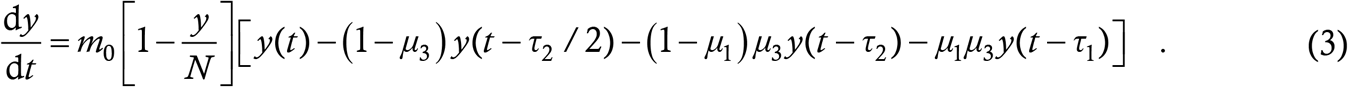

This is basically the DDE (1) from Ref. [2] with a logistic correction. We claim that this equation and variants which account for more sophisticated contact tracing measures, intentional quarantine violations etc. act as the overall governing equations for the spread of COVID-19.

Although it is possible to level criticisms against (3), for example that it does not account for the fraction of the susceptible population in temporary quarantine, that the logistic term tacitly assumes *y* to be constant during the intervals *τ*_1_ and *τ*_2_ etc, such criticisms can be aimed at any lumped parameter differential equation model. As we have already explained in Ref. [2], (1) is a very reasonable equation which captures the salient features of COVID-19 spread without being excessively complex. On top of that, the logistic correction is a very plausible quantitative description of the reduction in transmissibility arising on account of existing infections. The growth in new infections is automatically zero if *y* = *N*; at the same time, *y* = const. for any constant remains a solution which allows the outbreak to be over without reaching this maximum infection level. Note that the logistic term is a herd immunity effect, since it is a transmissibility reduction occurring as a result of increased prevalence of infected people.

The mathematical analysis of the logistic DDE (3) as a new type of nonlinear DDE will very likely be a fascinating exercise, but we leave this exploration for a future study. In this Article, we shall only solve the equations numerically, using the routine we have already written [1], [2].

### §5 DATA FITS FOR HOTSPOT REGIONS

In this Section we perform data fits for New York State and Italy to our model, to demonstrate its credibility. The details of the fitting process can become very boring reading even though they are very necessary, so we first give a quick summary. As we have already mentioned, the recorded case histories in these regions especially in the pre-peak regime have little credibility on account of the scarcity of testing, and the high numbers of asymptomatic cases going undetected. Hence, we use the death histories to extrapolate the actual case histories and then attempt a fit of the data to (3) using suitable parameter values. For both regions, we consider only the period of full lockdown since *m*_0_ is expected to be approximately constant then. We find good fits in both regions for the following approximate parameter values : *m*_0_ slightly less than 1/4, *μ*_1_ approximately 75 percent, *μ*_3_approximately 80 percent, and *N* approximately 80 lakhs which corresponds to a sizeable fraction of the population of both the New York Metropolitan area and Lombardy, Italy, where the maximum cases are concentrated. In the absence of detailed data from local medical and government authorities, the parameter space is over-rich which makes the fits non-unique; nevertheless we believe that good fits for physically plausible values of the individual parameters lend credence to our model. The rest of this Subsection is just amplification of these points, and can be skipped without loss of continuity.

For New York State, we have collected the daily reported case and death data over the period 01 March to 27 May 2020. We now assume that the mortality rate is constant and that the time to death is constant at 20 days. Then, we ignore the reported case counts and assume that the true case counts are the death counts multiplied by some number *α*, and shifted back in time by 20 days. Here, *α* is the inverse of the mortality rate. We treat it as unknown, to be determined during fitting. Due to limited testing capacity, the assumed actual case counts during the early part of spread have no relation to the reported counts. We then assume that, when the reported peak in case counts is reached, all symptomatic cases are being accounted for. This is because, by our hypothesis, the climb in cases prior to the peak is only an artefact of the limited testing capacity, and in such a situation, symptomatic cases will be given precedence in testing. The reported peak will be attained just when all new symptomatic cases can be accounted for, and thereafter the increased testing capacity can be used to start detecting asymptomatics also. This assumption enables us to determine *μ*_1_ given the value of *α*.

Full lockdown was imposed in New York State on 22 March 2020, which corresponds to the 22^nd^ day of the data. Since *m*_0_ will be fluctuating wildly prior to the lockdown, we start running (3) after the lockdown. The DDE needs to be seeded with an initial function which runs for the maximum delay involved in the problem, which is 7 days. Accordingly, we treat the assumed actual case counts of the 22^nd^ to the 29^th^ day as the seeding function, joining the discrete available data points by straight lines to create a continuous function. We then adjust the parameters of (3) so that the evolution of *y* in the subsequent period gives a good fit to the assumed actual case counts. Since these are derived from the death counts, they are available only upto a time of 20 days before the last day of data i.e. upto 07 May.

We find a good fit for the values *α* = 100 (mortality rate 1 percent), *μ*_1_ = 0.7334 (which follows from the prescription above), *μ*_3_ = 0.75 (when a region is overwhelmed with cases, contact tracing will not capture too many of them), *m*_0_ = 0.23 (pretty low, as appropriate for a full lockdown) and *N* = 8×10^6^. The plots are given below where the blue lines are derived from the model while the green bars denote the assumed actual case histories.

For Italy, the data starts from 20 February 2020. The lockdown was imposed on 09 March so we again start seeding from the 21^st^ day. The process is identical to what we did for New York. We again fix *α* = 100, which leads to *μ*_1_ = 0.8183. We assume *μ*_3_ = 0.9 since New York State is expected to be more efficient than Italy at contact tracing and also since Italy was the first country to be hit by this virus so there was no warning and no procedure in place for what should be done. Thereafter, *m*_0_ = 0.22 and *N* = 8.6×10^7^ produce the fits we give below.

As we have already mentioned in the summary, the parameter space is multi-dimensional and without insider knowledge (for example of how many patients are falling sick after being detected from a contact tracing drive) there will very likely be excellent fits in an entire region of that space contiguous to the point where we have achieved ours. Hence, the fits should not be taken as proof that the mortality rate is indeed 1 percent or that *m*_0_ during a lockdown is exactly 0.22. However, they should be indicative of the fact that the parameter values are in the right ballpark. Moreover, we get encouragement from two facts. One is that the well-fitting values of *m*_0_ naturally came out very close in the two regions – we have not doctored anything to make this work. The second is that the values of *N* came out to 8 million in New York State and 8.6 million in Italy. In New York State, almost all the cases are concentrated in New York City and the adjoining area. The population of this area is approximately 20 million, among which 8 million seems a reasonable estimate of the susceptible and at-large fraction. In Italy, about one third of the cases are coming from Lombardy so this part might be given an effective *N* of about 2.9 million. This is again a reasonable fraction of the total 10 million population of Lombardy.

Due to all the variables involved in fitting, we have desisted from attempting or presenting fits of more regions. Rather, armed with confidence from these two fits, we now illustrate some general properties of the solutions of our model (3).

### §6 SOLUTIONS OF THE LOGISTIC DDE (3)

We first derive the reproduction number *R* from (3). For the linear equation (1), *R* follows from the stability of a constant solution [2]. For the nonlinear equation (3), if we want the stability of the solution *y* = *y*\*, then we must first linearize about this point and then carry through the procedure detailed in Ref. [2]. This yields the same expression as (2) with *m*_0_ replaced by *m*_0_ (1 − *y*\*/*N*). Now, since *y*\* is arbitrary, we can replace it by *y* and write

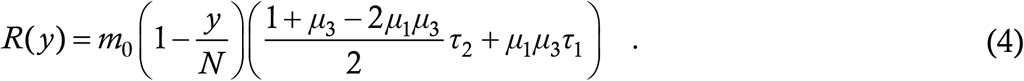

We would like to highlight that the definition of the reproduction number, and not just the starting value *R*_0_, comes out naturally from our DDE model. The extraction of *R*_0_ from an S-I-R style ordinary differential equation (ODE) model has been demonstrated by Diekmann et. al. [20]. These authors first linearize the system about the disease-free equilibrium and then isolate the transmission and transition matrices to obtain *R*_0_. In the ODE formulation, *R*_0_ < 1 corresponds to stability of the disease-free equilibrium while *R*_0_ > 1 corresponds to instability of the said equilibrium. We note that in reality, even when *R*_0_ < 1, introduction of the disease into society does lead to an increase of cases before the epidemic dies out – the system does not return to the disease-free state. This fact is captured by the DDE model. Moreover, the calculation of the instantaneous reproduction number at an arbitrary point of the disease evolution appears to be impossible from the ODE formalism since it requires linearization about a fixed point, and every disease state is not a fixed point of the system. Any constant solution is however an equilibrium of the DDE (3).

Having obtained the reproduction number, we now demonstrate various classes of solutions of (3). To this end, we consider a notional city having an initial susceptible population of 3 lakhs, fraction of asymptomatic carriers 80 percent and an initial condition of zero cases to start with and 100 cases/ day for the first seven days. We first consider Notional City A having the parameter values *m*_0_ = 0.23 and *μ*_3_ = 1/2, corresponding to moderately effective contact tracing in a hard lockdown. This gives a starting *R* of 0.886. Predictably enough, the epidemic burns itself out, as shown below. In this and subsequent plots in this Section, we show three things in the same plot – the cumulative case count *y* (*t*) as a blue line, its derivative *y*(*t*) as a green line and the weekly increments in cases, or the “epi-curve”, as a grey bar chart. For better visibility we have scaled down the weekly increments by a factor of 7. We report the rates on the left hand side *y*-axis and the cumulative cases on the right hand side *y*-axis.

We can see that the rate decreases monotonically; the maximum rate is 100 cases/day, during the seeding period.

In Notional City B, everything remains the same except that the contact tracing is less effective : *μ*_3_ (the escape fraction) is 75 percent. This gives a starting *R* of 1.16, which reduces to unity at *y* = 40500 cases.

We can see that the outbreak takes off quickly, entering exponential growth right after being released. As *y* increases, *R* gradually reduces so the growth slows down until it peaks when the case count is about 39,000. This is very close to the point where our analytical *R* crosses the unity threshold, and from that point onwards, the epidemic proceeds along a decelerating path to self-burnout. The overall progression of the disease is extremely protracted with the time to the end being almost double of the previous case. However, the small size of the peak should prevent overwhelming of City B’s healthcare facilities and thus avoid unnecessary deaths.

We now consider Notional City C which has the same parameters as City B except that *m*_0_ is 0.5. This means that the state of lockdown is replaced by a state of much milder restriction on mobility. The starting *R* is above 2.5 and it comes down to unity only at 1.8 lakh infections.

This time, the disease explodes like an atomic bomb, tearing through almost the entire population in a very short period and presumably leaving behind a trail of dead bodies in its wake.

Notional City D is a combination of B and C. This city starts off with *m*_0_ = 0.5 like City C but then realizes its folly and cuts down instantaneously to *m*_0_= 0.23 like City B when the number of cases becomes 40,000.

The sudden lockdown throws boron onto the atom bomb before it can reach full detonation power. The ultimate upshot is about 70 percent more cases than City B but a significantly shorter time to the end of the outbreak, which can offset the higher case cost, depending on the urgency of the external economic and other conditions. However, the peak rate of 12,920 cases/day is still very high and likely to overstress all but the world’s most advanced healthcare systems.

Now we consider two cities which are trying to reopen after a lockdown. In both these cities, we start things off with the parameter values of the self-burnout City A. Then, the city reopens on the 80^th^ day by increasing *m*_0_ from 0.23 to 0.5, and simultaneously decreasing *μ*_3_. In City E, *μ*_3_ = 0.1 after reopening i.e. 90 percent of all cases get detected and isolated through contact tracing when the reopening is achieved. This generates an *R* of 0.985 on the 80^th^ day; here is the time trace of evolution.

The sudden increase in *R* at the reopening increases the time to burnout. Indeed, the time taken is double that of City A. However, this excess time comes as the flip side of much higher social mobility, and there is very little extra case cost. Thus, the lockdown has been extremely effective, allowing retardation of the disease while the contact tracing capacity was being built up. Another important feature of the plot is that, since *R* < 1 throughout, there is no increase in the case rate at any stage.

City F is almost the same as City E, but for the fact that the post-reopening *μ*_3_ is 0.2; 80 instead of 90 percent of cases are captured through contact tracing. This raises the *R* at reopening to 1.22, which comes down to unity only at about 53,000 infections. The time trace of evolution is below.

This is a quintessential reopening disaster – the contact tracing after the reopening was not sufficient to sustain the higher mobility level, and the cases ballooned in a second wave after the 80^th^ day, just as in City B. The 80 days of full lockdown at the beginning ended up being completely futile and it would have been better to go with the post-lockdown parameter values right from the start.

Finally, we note that if the susceptible population *N* of each city is varied while keeping all other parameters constant, then the graph heights scale almost in exact proportion to *N*, and the epidemic durations vary by only minuscule amounts.

### §7 TESTING-INDUCED DISTORTION

In this Section we see how changes in testing capacity can result in distortion and misrepresentation of a region’s epidemiological curve. We consider the two hotspot regions of Maharashtra and Delhi. For each region we harvest from Ref. [3] a dataset consisting of the number of tests done on each day between 18 April and 25 May, as well as the number of cases found on each day within this interval. We present plots containing this information, as well as the ratio of the number of tests to the number of cases for each day.

Before discussing these plots, we mention what an ideal testing vs. cases curve looks like. Ideally, a region has huge testing capacity to begin with – say one lakh tests per day in a city. In the early phases of the outbreak, the case rate is low, so the test-to-case ratio is very high. As time evolves, the testing capacity remains constant but cases increase exponentially, so the ratio dips rapidly. Then, beyond the peak, the ratio increases again as the cases thin out. More practically, there will also be some increase in testing capacity with time, but due to the high capacity at the start, that increase will be much slower and smaller than the increase in cases. The ratio should still show a clearly descending profile before the peak and an ascending profile after.

Now let us compare this with reality. In Maharashtra, the tests/cases ratio is approximately constant at around 12 between days 1 to 16 (18 April to 03 May) and approximately constant at around 6 between days 17 to 38 (04 to 25 May). Indeed, the mean of the ratio during this latter interval is 6.04 and the standard deviation is 1.51, which decreases to 1.06 if we eliminate the single lowest and highest outlier values. This suggests that the ratio is very nearly constant between days 17 to 38, which is a significant deviation from ideal behaviour. During this time, the case rate increased from about 1200 to more than 2500 cases/day, representing a more than doubling of the rate. However, the fact that the testing rate doubled as well casts doubt on the veracity of these figures – we suspect that if there had been more tests available earlier, then Maharashtra would have found more cases earlier. Delhi by contrast does show a decrease in the ratio, especially beyond the 20^th^ day. The ratios themselves are also much higher than in Maharashtra which suggests better testing capacity. This gives more credence to the reported case histories of Delhi.

As another example of the influence of testing on reported case histories, consider the daily case increases in India over the period of 29 May to 02 June. They are 8138, 8364, 8789, 7723 and 8815 respectively.

Now, the number of tests done on these days are 1,21,702, 1,27,761, 1,25,428, 1,00,180 and 1,28,868 respectively. The one day that there is a lower case count, there is also a lower test count. We cannot help but wonder that if India were suddenly to be gifted with say one crore testing kits, then how many cases would we find.

There are two more issues which lead to distortions in the reported case counts. The first is that mildly symptomatic people sometimes do not report for testing, for fear of contracting the disease from the testing centre if they happen to not have it already. While we are not aware of any incident where corona has actually spread from a testing facility, the fear nevertheless remains in public mind. The second issue is the reliability of the test itself. When testing of persons without symptoms is carried out as a result of contact tracing, it can happen that the test ends up getting done during the disease incubation period. Unfortunately, real time RT-PCR test, which is the most widely used protocol today, has very high false negative ratio [21], [22], ranging from certain failure on the day of exposure to 25 percent probability of failure even at the height of symptomaticity in patients (antibody test is useless for diagnostic purpose and we are not aware of the statistics for TrueNat and CBNAAT tests). RT-PCR will almost certainly fail to catch many asymptomatic carriers who get tested prematurely, find negative and then don’t seek testing any longer since they don’t develop symptoms. To reduce this risk, the Indian Council of Medical Research (ICMR) has stipulated that potential cases arising from contact tracing must be tested between 5 and 10 days from the suspected exposure date. Even with this precaution, distortion of the actual figures is bound to occur.

### §8 PARTIAL HERD IMMUNITY AND GLOBAL SPREADING DYNAMICS

In this Section, we explain the concept of partial herd immunity. Notional City B from Section 6 is the best demonstrator of this. Here, the epidemic was started off in unfavourable conditions and grew exponentially as we would expect. However, as it grew, herd immunity started kicking in, to an extent that the disease entered a stable zone (*R* < 1) when only 13.5 percent of the total susceptible population was infected. From this point onwards, it got extinguished in time like the self-burnout City A, infecting another approximately 13 percent people in the process. Thus, in this example, herd immunity worked in concert with public health measures to achieve a halt to the epidemic with only 26 percent of the population infected, which is significantly lower than the conventional herd immunity threshold of 70-90 percent [23]. This is what we call partial herd immunity. Britton et. al. [24] and Peterson et. al. [25] both find lower herd immunity thresholds than the conventional value; we believe that our work explains these findings. Note that with the social restrictions off, partial herd immunity does not work any longer, as in City C. Here, the disease goes all the way upto 90 percent infection. The maximum case rate increases by a factor of 25 relative to City B, very likely straining the medical resources of City C multiple times beyond their limits.

We have observed an interesting fact about cities of Type B which start off in the red but enter the green after a short time. We find that if *R* = 1 occurs at a population fraction which is significantly less than 1/2, then the cumulative case count at the end of the epidemic is very close to double the value at which *R* = 1 is breached. We suspect that we shall find exactly double in some limit, and we designate this as one of the questions to address in a mathematical treatment of (3).

One more feature of (3) is that it does not have a solution *y* which is linear in time, except unless *R* = 1 identically, which is a marginal case. Similarly, none of the other epidemiological models in vogue also admits a solution which is proportional to *t*, i.e. where the case rate is a constant. Yet, we have been seeing linear case profile in multiple countries and regions such as Canada, Sweden, California and Maharashtra (days 1 to 14 in Fig. 8) – still more examples can be found in Verma et. al. [26]. One factor which can contribute to explaining these growths is testing limitations as discussed in the previous Section. A second – and completely different – explanation has been given by Thurner et. al. [27] in a very recent study. They have modelled the spreading of COVID as a stochastic state-change of each lattice site (person) in a small-world network, and for some properties of the network they have found solutions which grow linearly until they stop abruptly. On the whole however, their solutions are in excellent agreement with what we have got for the Notional Cities A to D – where they find progression to herd immunity, so do we; where they have linear growth till an abrupt stop, we have a gentle S-shape to a smooth stop. After a point, a reality of this nature (unlike a physical law) is not governed by mathematics – any model is at best a representation and a simplification.

**Figure 8:**
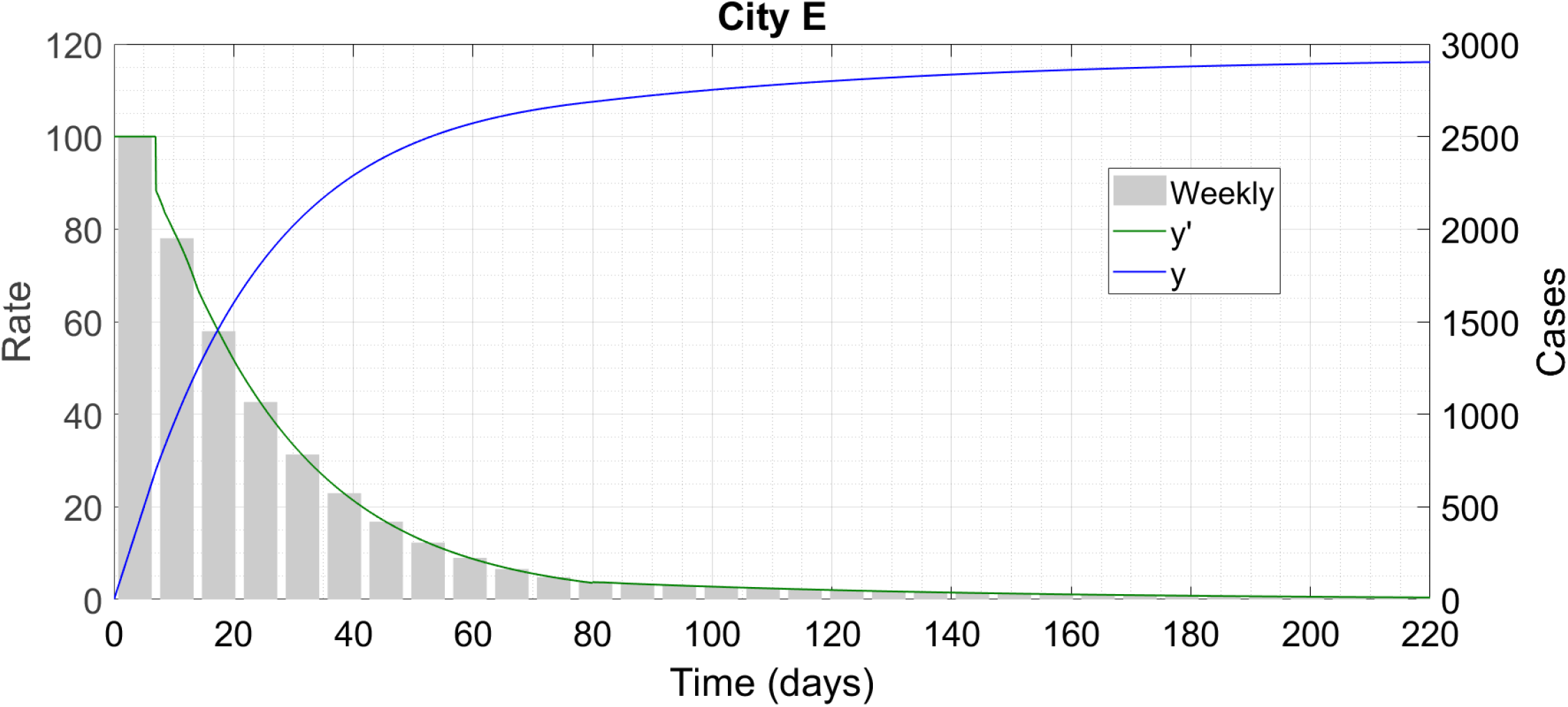
Like City A, City E also burns the epidemic out.

**Figure 9:**
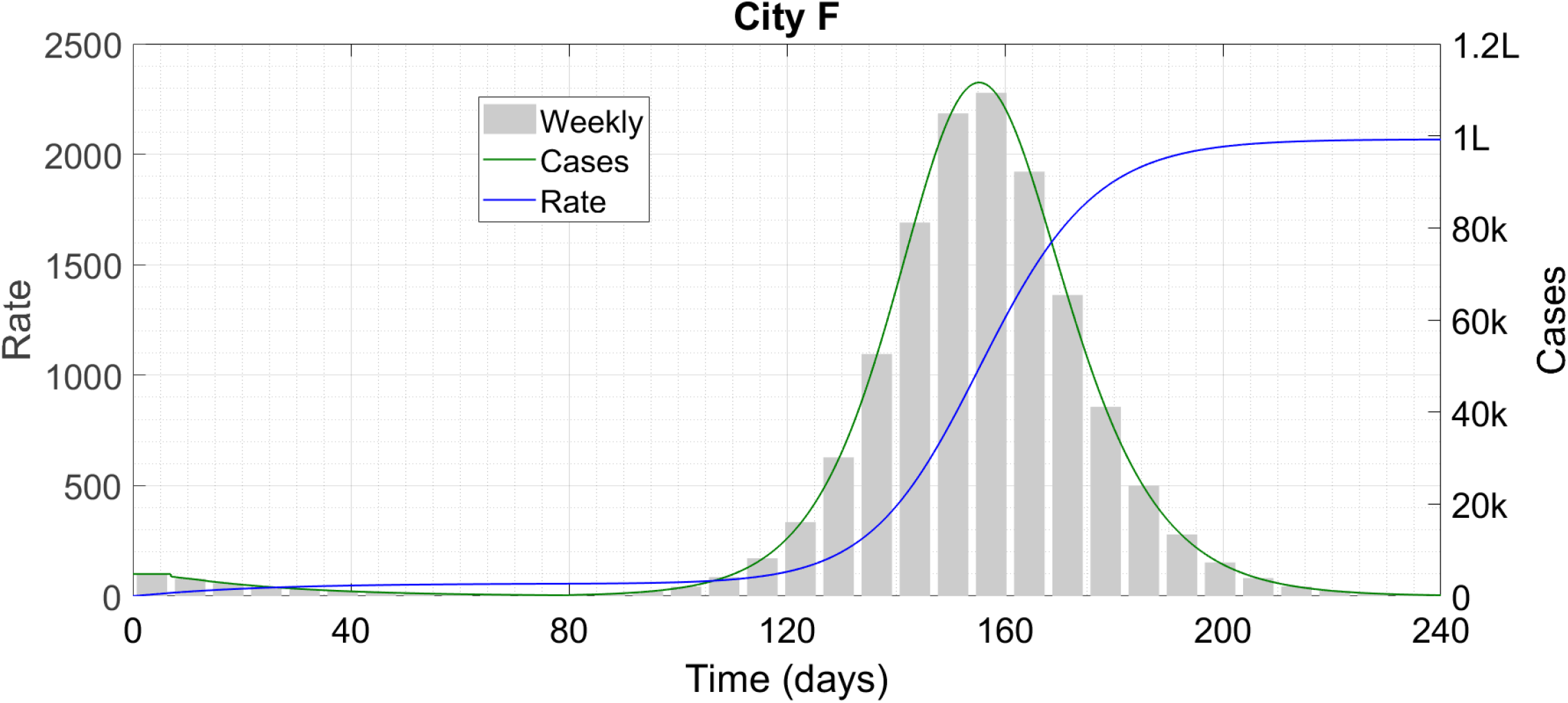
After reopening, City F becomes similar to City B.

**Figure 10:**
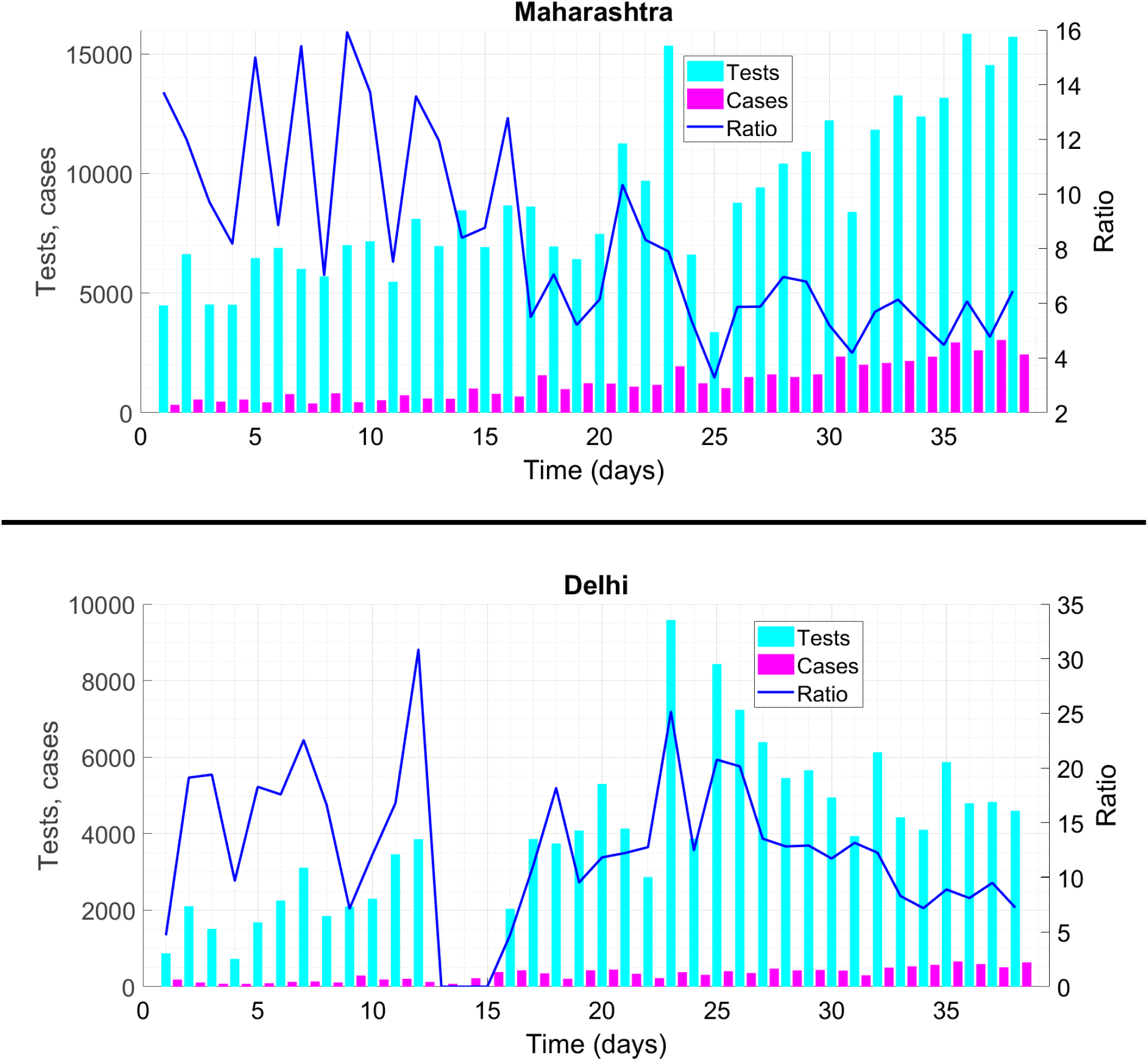
Tests done per day (cyan bars), cases recorded per day (magenta bars) and ratio of these two quantities (blue line) for Maharashtra (upper panel) and Delhi (lower panel).

**Figure 11:**
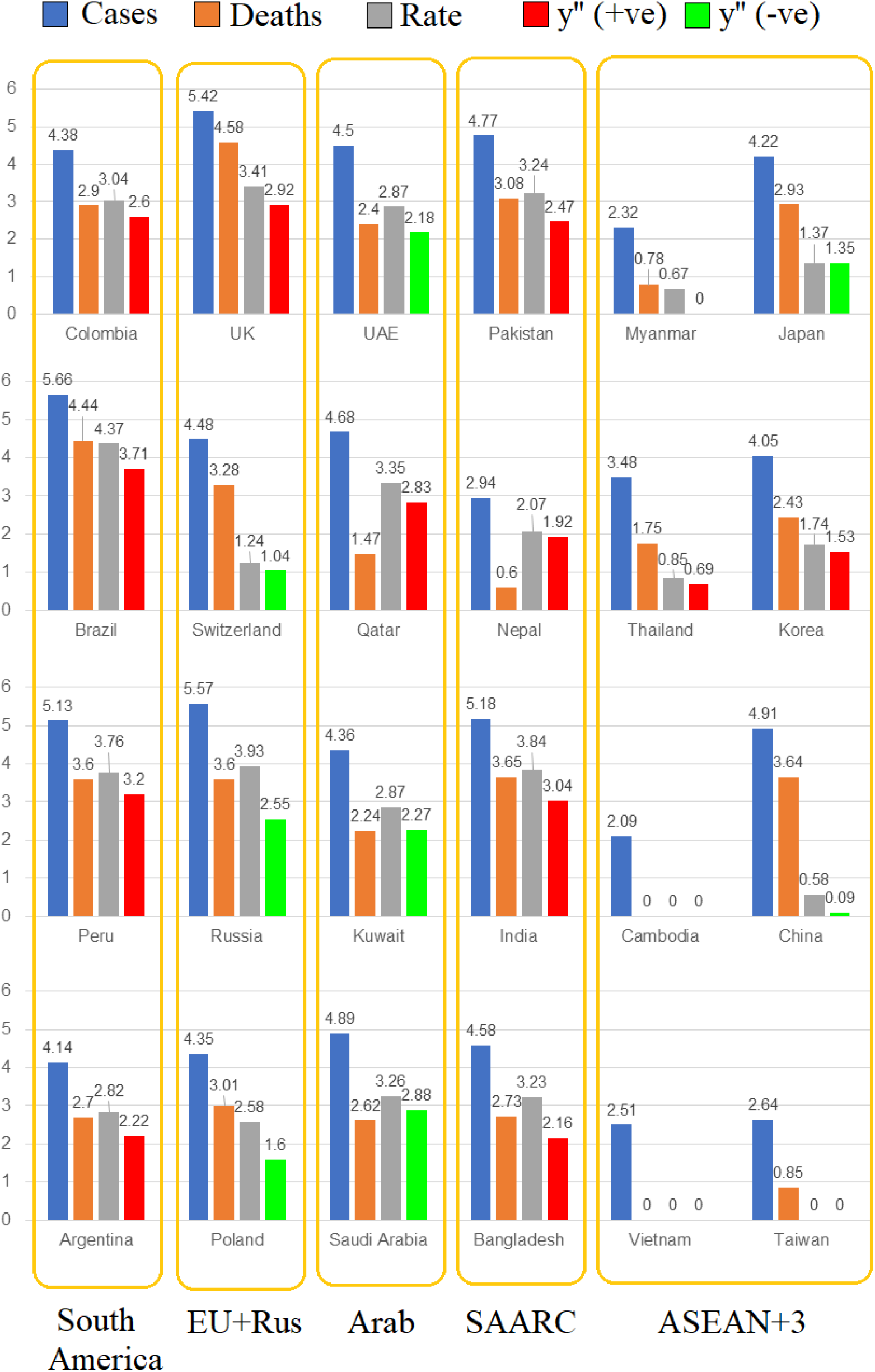
Corona statistics in various regions of the world. Note that the y-axis is logarithmically scaled so 4.38 for Colombia cases means that Colombia has 10^4.38^ = 24104 cases. For obvious reasons we do not take a logarithm of a zero value but display it as zero.

The global spread of COVID-19 appears extremely complex and individual cities, districts or counties (perhaps even neighbourhoods) are behaving like Notional Cities A, B and D (we believe that nobody is so negligent as to go with strategy C), and the resulting country- or statewide response is a sum of these localized effects. Mumbai in Maharashtra and Los Angeles in California for example are very likely cities of Type B since they locked down early but could not do the requisite contact tracing and gradually spun out of control. We expect that unless mobility restrictions are lifted rashly, they will continue on a not-so-slow burn trajectory until partial herd immunity is reached, at which point they will start coming out of it. New York City on the other hand is likely of Type D since it remained open during the initial (mostly undetected) explosion of cases. Consistent with our analysis, the City flared up dramatically, reaching high overall case counts but it is now out of the woods and should be able to start the reopening process within the next week or so without seeing a second uptick in cases. Finally, there are countries such as Austria, Australia, New Zealand and Israel, and states such as Kerala in India, which are close to self-burnout of the epidemic and must be composed entirely of Type A cities.

We believe that equations having the basic structure (3) with different parameter values to account for the different lockdown and contact tracing situations, combined with the anomalies arising from testing limitations, can explain the case histories of Coronavirus all over the world. Naturally, the model (3) can also be used for predictive purposes. Once a case history is obtained, auxiliary parameters such as required hospitalization capacity, death count, PPE requirements etc can also be calculated without too much difficulty. The simple structure of (3), relative to models such as SIDARTHE [28], SEIR-QDPA [29], extended SEIR [30] and network based model [31], make it easier to work with while producing equally, or more, detailed results. It is also interesting that the governing equation is in some sense a small variation on the original logistic equation, which has frequently been used for modeling corona dynamics [32]–[34]. Whereas these works often appear overly simplified, the addition of the delay enables us to encapsulate a lot of the realistic phenomena associated with the transmission of this virus.

We do not however declare (3) itself to be the most accurate possible representation of Coronavirus dynamics. The following refinements are possible for example :

- Accounting for the latency and incubation period separately instead of in one lumped delay *τ*_2_
- Accounting for bi-directional contact tracing and not just forward from symptomatic patients
- Accounting for the time taken in contact tracing and for intentional violations of quarantine by irresponsible citizens
- Accounting for temporary quarantine of healthy people on suspicion of having been exposed to COVID
- Accounting for multiple population categories with different degrees of susceptibility and risk

Here we have not attempted these refinements since they will destroy the simple structure of (3) and introduce a plethora of additional parameters of whose values we are currently ignorant. But, whatever the modifications made, the basic structure of a logistic term multiplying some *y* − *y*_delayed_terms should remain intact, and this can help us to understand and forecast the dynamics of the virus all over the world.

## 3 IMPLICATIONS OF THE MATHEMATICAL RESULTS

In this Part we shall discuss some broader implications of our work.

### §9 REOPENING CONSIDERATIONS, ROLE OF TESTING

The unemployment situation generated as a result of lockdowns is currently forcing countries and states to partially reopen their economies even though many of them have not yet got the virus under control. The reopening is easiest in Type A regions where cases have slowed down to a trickle. By definition, contact tracing was efficient in these regions to begin with; moreover, the slow rate of growth means that now it is even more effective. With every new case being detected, swift isolation of all potential secondary, tertiary and maybe even quarternary cases, both forward and backward, should prove possible while the rest of the economy functions in a relatively uninhibited way. Of course, separation minima, sanitization, masks and every restriction of that kind should remain strictly in force as we have already discussed in Ref. [2]. Even one mass transmission event can restart an exponential growth regime and force a rollback to a fully locked-down state.

Reopening beyond a skeletal level is impossible in Type B regions which are still in the ascending phase. The ascent implies that contact tracing is already inadequate, and on top of that if mobility increases then the region might turn into Type C, overstress healthcare systems, and become a massacre. An ascending B-City has little option other than to contact trace as hard as possible and wait for partial herd immunity to kick in. Only when that happens and the cases slow down on their own can it consider a more extensive reopening like a Type A region.

Testing is an important part of the epidemic management process no doubt since it enables the authorities to get an accurate description of the spread of the disease. As we have already discussed, limited testing capacity is giving us a partial or distorted picture in many regions. There is a widespread media perception that extensive testing is one of the prerequisites for any kind of reopening process [35], [36]. Much criticism has also been levelled at certain countries for having inadequate testing programs (we shall further elaborate the blame aspects later). However, we would like to emphasize that testing is as of yet a diagnostic tool and not a preventive one. Currently, it can show us how the disease is behaving but cannot slow its spread in any way. Test-induced slowing can come only when the capacity expands to such a level as to be able to preventively test potential super-spreaders such as grocers and food workers every single day.

That said, during reopening it is vital to get a true picture of the disease evolution so that we can gauge the effect of any relaxation of restrictions – whether it keeps the outbreak under control as in City E or brings about the beginnings of a second wave as in City F. Such beginnings are heralded by a rise in the case rate – as we saw, there was no such rise in City E even though *R* increased after the reopening. If the rise takes place, the relaxation must immediately be rolled back to avert the disaster. Hence, during reopening, the testing capacity must be high enough to detect such incipient rises. As per China’s state media reports, with an aim to reopen the economy, the city of Wuhan conducted 6 million tests in one week; we present this fact without discussion or comment.

A second reason why testing is still not all that it could have been is the high false-negative rate during the initial stages of infection, as already mentioned in §7. Suppose a contact tracing drive identifies Mr X as a potential case, having been exposed to a known case yesterday. Then, it can be that Mr X contracts the virus ten days from now, in which situation he will report negative if tested today or tomorrow, but will still amount to a spreading risk ten days later if he is at large then. This also means that secondary contact tracing, i.e. finding Mr. X’s contacts, must go ahead irrespective of his test results. Indeed, the medical authorities are well aware of this loophole. We will quote verbatim some sections of the public announcement arising from a confirmed case of an employee at a grocery store in Ithaca. The case was detected on 14 May. Immediately, customers who shopped at the store on 08, 09 and 11 May were advised the following :

- It is recommended that you get tested at the Cayuga Health Sampling Site [the testing facility in Ithaca].
- Self-quarantine in your home for 14 days from the last date you shopped.
- If you seek testing and the result is negative, continue to self-quarantine and monitor yourself for the full 14 days from the last time you shopped. If you become symptomatic, seek testing again.

The third point – self-quarantine irrespective of the test result – makes it amply clear that it’s the quarantine and not the testing which is actually going to stop the spread of the disease. We can compare the spreading and testing situation to an electric locomotive which has potentially under-performing traction motors and faulty ammeters. Since the ammeter is defective, the loco pilot doesn’t get to know how much current the motors are drawing, whether the motors are in good condition or otherwise, and how rapidly he can increase the throttle setting and accelerate the train. However, we have seen some loco pilots who, despite a dead ammeter, figure out the motor condition by instinct during the first few minutes of run, and then drive optimally within the motor’s limits. If the motor turns out to be in poor condition, then repairing the ammeter alone will not result in a good run – the motor needs to be replaced as well. However, it is also true that to systematically extract the maximum performance from a locomotive with highly rated motors, we do need fully functional ammeters as well [37].

In a recent series of discussions pertaining to the reopening of Cornell University Ithaca campus and the possibility of in-person classes in the Fall Semester (2020-21/I), some people were voicing the opinion that in-person classes might be possible if testing capacity is adequate. We would beg to differ. Firstly, it is impossible that there will ever be a testing capacity high enough to do a real time RT-PCR every day on every single member of a 25,000-strong community. But let’s say we use cluster testing [38] to collectively test all the members of every course once a day. Suppose that one day, one course tests positive. Then, all students as well as faculty of the course will have to be immediately quarantined and individually tested. Instruction will at once stop, and the testing process itself might become nightmarish if the course involved is a large one. Moreover, student cases who falsely test negative on account of the testing limitations discussed above might not be convinced to remain in quarantine nonetheless. The only way that in-person classes can resume, in our opinion, is if the following conditions all hold true :

- Ithaca locally becomes free of virus (no new cases for at least 3 weeks)
- All students arriving from virus-prone regions are institutionally quarantined for 15-20 days upon arrival
- All cases among the external arrivals are detected and recovered
- The perimeter of the City of Ithaca (or of a larger contiguous virus-free area) is sealed so that residents of the safe zone cannot venture out and come back with virus

While we have expressed this in terms of Cornell and Ithaca, these considerations remain valid for any university anywhere in the world which is considering reopening. If all the above conditions cannot be satisfied, we feel that Universities should remain in the online teaching mode until the pandemic is over.

In the rest of this Section, we mention the reopening guidelines stated by authorities in USA, and show that they are by and large in agreement with what we discussed above. If you would like to skip it (for example you are not located in USA), please go to the next Section. Separation minima, wearing of masks and similar basic steps need to continue during reopening. CDC has issued certain recommendations for effective ways of commuting. Extra attention is required in high-risk areas like restaurants, moderately occupied offices, theatres, supermarkets, hair-dressers etc and the public should be aware of the necessary precautions. The US Chamber of Commerce has given out state by state reopening guides for small businesses which are mandated to be followed across the US. Continued following of federal, state, tribal, territorial and local recommendations is of paramount importance.

Prior to resuming work, all workplaces should have a carefully chartered exposure control, mitigation and recovery plan. Although essential guidance is specific for each business, there are certain measures that can be generally adopted across all workplaces.

1. Reopening in phases – The US government has laid down guidelines to open the country in 3 phases. First phase involves continuation of vulnerable individuals to remain at home. When in public, people are expected to wear masks, have maximum physical separation, avoid places with more than 10 people and limit non-essential travel. Second phase allows gatherings of 50 people, some non-essential travel and reopening of schools. Third phase involves relaxation of restrictions, permitting vulnerable populations to operate.
2. Defining new metrics – Post-corona world will witness some significant changes in regulatory controls, and behavioural drift in personal and professional spheres. Cleanliness standards, safety standards, infection prevention practices with regular monitoring and inspection for its assurance are some of the new terms that will have to be a part of a daily life of the people for at least the next few months.
3. Organizational changes – To help essential operations to function, companies and organizations will have to be prepared with advanced IT systems (in case of continuation of remote working), supply of PPE, setting up travel facilities to avoid public transport, providing behavioural health services, and leave no stone unturned in overcoming biological, physical, and emotional challenges.

We can see that the above guidelines are broadly conformal to our model predictions.

### §10 METHODS OF CONTACT TRACING

As we have already mentioned, contact tracing is probably the single most important factor in determining the progression of COVID-19 in a region. In the model equations (1) and (3), we have assumed instantaneous contact tracing starting from symptomatic cases. Multi-level contact tracing will very likely be required to ensure that 75 percent or more of cases can be quarantined before they turn transmissible. The faster the contact tracing takes place, the better – if it takes a time *τ* to isolate contacts then that will add on to *τ*_2_/2 in (3); as we can see from (4), the more delay terms we have, the higher *R* becomes. Moreover, our model does not account for backward contact tracing. In practice however, a sufficiently high level of detection might not be possible to achieve with forward contact tracing alone.

As much as it is important, contact tracing is also one of the trickiest aspects to handle since it can interfere with people’s privacy. In classical contact tracing, human tracers talk to the confirmed cases and track down their movements as well as the persons they interacted with over the past couple of days. This method has worked well in Ithaca and in Kerala. While it is the least invasive of privacy, it is also the most unreliable since people might not remember their movements or their interactions correctly. The time taken in this method is also the maximum. A more sophisticated variant of this supplements human testimony with CCTV footage and credit/debit card transaction histories – this approach is possible only in countries such as USA where card usage predominates over cash. The most sophisticated contact tracing algorithms use artificial intelligence together with location-tracking mobile devices and apps – while they are quick and fool-proof, they automatically raise issues of privacy and security. For example, the TraceTogether app in Singapore, which worked very well during the initial phases of the outbreak, has not found popularity with many users [39]. Similarly, India’s Aarogya Setu has also raised privacy concerns [40]. Americans too have expressed their aversion to using contact tracing apps in a recent poll, with only 43 percent of people saying that they trusted companies like Google or Apple with their data.

Recently, there have been reports saying that American legislators have outlined a plan to regulate the process of digital contact tracing. States have been instructed to develop their own apps (for example – Healthy Together in Utah, Care19 in North Dakota etc). Companies such as Deliotte have come up with ABTrace Together application which has been downloaded more than 1,00,000 times and has undergone a privacy impact assessment to get a green signal for wide application in its intended purpose. The statement issued by this application also acknowledges that it is in compliance with the Health Information and Freedom of Information and Protection of Privacy Act.

### §11 ENSURING SOCIAL COMPLIANCE – A BEHAVIOURAL PERSPECTIVE

As the epidemic drags on and on, the continued restrictions on social activity are becoming more and more unbearable. There is an increasing tendency, especially among younger people who are much less at risk of serious symptoms, to violate the restrictions and spread the disease through irresponsible actions. However, as we saw in §6 for City F, a rise in violatory behaviour can completely nullify the effects of lockdown over the past few weeks or months. Here we discuss how public health professionals and policy makers can resort to behaviour/psychological theories to ensure compliance among the common people. The most widely used model is Health Belief Model which has been used successfully in addressing public health challenges. We briefly discuss the utility of this model in the current situation.

Health belief model is a theoretical model which hypothesizes that interventions will be most effective if they target key factors that influence health behaviours such as perceived susceptibility, perceived severity, perceived benefits, perceived barriers to action and exposure to factors that prompt action and self-efficacy. In general, this model can be used to design short and long term interventions. The prime components of this model which are relevant in the current scenario can be outlined as follows.

1. Conducting a health need assessment to determine the target population – The best example is the demarcation of zones in India depending on the level of risk. Red zone is highest risk, orange zone is average risk and green zone translates into no cases since last 21 days. Classification is multifactorial, taking into account the incidence of cases, the doubling rate and the limit of testing and surveillance feedback to classify the districts.
2. Communicating the consequences involved with risky behaviours in a transparent manner – Central and state ministers as well as public health authorities are in constant communication with the masses.
3. Conveying information about the steps involved in performing the recommended action and focusing on the benefits to action – Famous celebrities, in addition to state and central governments, spread the messages explaining the required steps cogently and ensuring that it has the maximum reach, especially among social media-addicted millennials and similar populations.
4. Being open about the issues/barriers, identifying them at early stage and working toward resolution – Activating all sorts of helpline numbers, email addresses, personal offices etc to address any grievances around the topic.
5. Developing skills and providing assistance that encourages self-efficacy and possibility of positive behaviour change – Adequate arrangements for people from lower socio-economic strata, stable and trustworthy financial schemes for middle class, plan to support small business and a means to become a bridge between the affluent class and the needy class are some of the ways to foster positive behaviour change and develop natural trust.

Other than health belief model, some theories that can be useful are:

### Theory of Reasoned action

This theory implies that an individual’s behaviour is based on the outcomes which the individual expects as a result of such behaviour. In a practical scenario, if the health officials want the people to follow a particular trend, let us say based on our model, they need to reinforce the advantages of targeted behaviour and strategically address the barriers. For instance, to enforce separation minima even when it is apparently proving ineffective and the cases are increasing, they can use the examples of Cities B and C to convince the citizens that violations – and hence violators – can be responsible for thousands of excess deaths.

### Trans-theoretical Model

This model posits that any health behaviour change entails progress through six stages of change : precontemplation, contemplation, preparation, action, maintenance and termination. For instance, it was observed that in March, despite a rise in cases in New York City (NYC), people were not observing social restrictions the way they should have. Now, we can see that with passing time, the behaviour of the masses transforms according to the stages of this model :

#### Precontemplation

This is a stage where people are typically not cognizant of the fact that their behaviour is troublesome and may cause undesirable consequences. There is a long way to go before an actual behaviour change. This phase coincides with the commencement of cases in NYC.

#### Contemplation

Recognition of the behaviour as problematic begins to surface and a shift begins towards behaviour change. When the cases started being reported all over media and the major cause of spread began to surface, citizens started paying attention to their activities.

#### Preparation

People start taking small steps toward behaviour change like in our case, exhibiting hygienic practices and ensuring separation minima.

#### Action

This stage covers the phase where people have just changed their behaviour and have positive intention to maintain that approach. In this instance, people continue to practise social restrictions and hygiene positively.

#### Maintenance

This stage focuses on maintenance and continuity toward the adopted approach. Majority of people in NYC are exhibiting positive behaviour and maintaining it throughout the stages of reopening phases. This is vitally important to ensure that NYC stops at partial herd immunity like City D instead of blowing up again like City C.

#### Termination

There is lack of motivation to come back to the unhealthy behaviours and some sections of people across the country/world will continue practising good hygiene (though not social restrictions!) in our day-to-day lives.

### Social Ecological theory

This theory highlights multiple levels of influences that moulds the decision. In our case, let us say for example that the decision is to maintain sufficient physical separation once offices are opened up. To successfully follow this, there is a complex interplay between individual, relationship, community and societal factors that comes into action. Law enforcement authorities need to take this into consideration. A group of individuals when motivated by one another to follow the guidelines, builds a good connection within the society, and in turn there is a high probability to build a healthy network within a defined area. A negative interplay at different levels of motivation may in turn, prove disastrous and cause all efforts go down the drain. A perfect illustration of this in the present condition is how various NGO’s are working in conjunction with public health authorities to bring about a change at an individual level by door-to-door campaigning. This propels the behaviour of even the most potentially recalcitrant population in the most desirable way i.e. wearing masks and gloves, adopting hand hygiene, being cognizant of symptoms arising in any member of the family and following quarantine rules in case of travel from other states.

### §12 A RACIAL FACTOR

Over the last couple of months, as corona has been spreading worldwide, we have been observing that some countries are having a disproportionately easier time of it than others. In the below bar chart, we consider several countries in different geographical regions and/or unions. For each country we show four things : (*a*) the total number of recorded cases upto 27 May, (*b*) the total number of deaths recorded upto 27 May, (*c*) the rate of new cases on 27 May and (*d*) the case rate on 27 May minus the rate on 20 May. For calculating the rates, we have performed a three-day running average twice centred on the day in question. The quantity (*d*) is basically the second derivative *y*’’ – if it is positive then the epidemic is in the pre-peak stage while if it is negative then it is in the post-peak stage. For visual clarity, we extract the common (base-10) logarithm of each quantity and display the resultant. For negative *y*’’, we take the logarithm of the absolute value; we show positive *y*’’ in red and negative in green.

It is a surprising observation that almost all countries in the easternmost union (ASEAN Plus Three) have got the COVID menace under control, which seems to indicate race as a factor of difference. Countries in this union are in a far better position with respect to the corona statistics than similar or more advanced countries in other regions and/or unions. From our equation (3), the factors which can lead to self-burnout are low *m*_0_ and low *μ*_3_. Now, it is impossible to believe that contact tracing is far more efficient in ASEAN+3 nations than in EU or SAARC nations. Hence, *m*_0_must be making the difference. Then, *m*_0_ has the two contributions from mobility and transmission probability. Social mobility, at least in some period of the disease management, was extremely low in almost all the countries considered in the above Figure, including India and Peru where the first and second derivatives are very high all the same. The only remaining factor is the transmission probability, which forces us to conclude that a close interaction between a sick and a healthy person in the less affected nations has a much lower risk of spread than a similar interaction in the other nations. This means that East Asians must be exhibiting significantly less susceptibility and/or transmissibility than people of other races. Only a few days back, a study has emerged [41] claiming that black people in New York City are more susceptible to infection than white people, so such racial factors might well be present here too. It had at one time been thought that the virus does not care about racial and national boundaries – that unfortunately doesn’t seem to be the case any longer.

While excellent contact tracing can account for the corona success of one or two among the ASEAN Plus Three countries – just as it has created isolated success stories within the European Union and the Arabian Peninsula – it cannot account for all of them. One more blow to the contact tracing hypothesis comes from the case cluster spawned by the premature reopening of all South Korea with the virus still around. Although this hasty behaviour did lead to new infections, the total count of those is just about 100 with fresh detections slowing down again. By contrast, an illegal but de facto “reopening” at just one market (Koyambedu Market) in Chennai (Tamil Nadu, India) has given rise to at least 2700 new cases [42]. These numbers lend further credence to the hypothesis that race is driving the corona success stories in East Asian countries. We currently have no idea regarding the aperture in the armour of this zoonotic virus which people of one race can penetrate but not another.

### §13 SOCIAL ATTITUDES AND BEHAVIOUR

In this Section we address another important issue related to the Coronavirus. This is that the widely heterogeneous case profiles in different regions have often led to “corona contests” among these regions. Far too often, the residents of better-off regions are seen heaping scorn on worse-hit regions. Leaders as well as the common people of regions suffering from virus come under fire from their more fortunate neighbours – the former for wrong decisions and the latter for non-compliance. Such scurrilous attacks appear everywhere from respectable media outlets to social media posts and individual telephonic conversations. We have selected a tiny handful of representative media articles, castigating the approaches of India, USA and Sweden, to show the breadth and vitriol of such commentary [43]–[49]. A feature common to almost all opinion pieces like this is that their authors do not have the slightest knowledge of the issues involved, either epidemiological or economic.

Before embarking on criticisms, we should note that policy decisions need to be taken in real time, as the situation evolves. The authorities do NOT have the benefit of hindsight to decide on their course of action. Since the virus is a new one, there is no precedent which can act as a model. Even among emerging infectious diseases, this latest one is particularly unpredictable, since minuscule changes in parameters can cause dramatic changes in the system’s behaviour (this might sound like chaos but it isn’t – can (3) or similar systems show chaos for some parameter values is a question we reserve for a later time). This phenomenon is best illustrated by the Notional Cities of §6. For example, to get from City A to B, all we did was increase by 50 percent the fraction of people who escaped the contact-tracers’ net. The result was a 30 times (not 30 percent!) increase in the total number of cases. Similarly, the difference between Cities B and D is an 11-day delay (recall that the first seven days in the plots are the seeding period, so they don’t count) in imposing the lockdown in D. 11 days out of a 200-plus-day run might not sound like a lot. But, that was enough to create tens of thousands of additional cases, risk overstressing healthcare systems and at the same time shorten the epidemic duration by a factor of three. To take yet another example, the difference between Cities E and F is 10 percent more cases failing to get quarantined in the latter city after reopening. That alone has caused 40 times more cases.

Further uncertainty comes from the fact that the parameter values are changing constantly. As we have mentioned in §2, the reported fraction of asymptomatic carriers has increased continuously over the last three months or so. Considering the sensitivity of this or any other model to parameter values, such changes can completely invalidate the results of a model as well as any decision which was made on their basis. The contact tracing parameter *μ*_3_, on which so much rests, is also extremely variable. Identifying potential exposures is much easier in a smaller city than a large or densely populated one. It is also more effective if the cases are mostly from the sophisticated social class who can use mobile phone contact tracing apps or otherwise keep (at least mental) records of their movements and of the people they interacted with. However, if there is an outbreak among the unsophisticated class, then even the most skilful contact tracer might run up against a wall of zero or false information. In such cases there are limited options that are left to the authorities to proceed in a conducive manner.

India went into lockdown on 25 March 2020. At that time, the official figures stated that there were only 571 cases, which made the decision appear premature to many people. Indeed, a seven-day delay of lockdown was suggested so that the migrant workers would have been able to return to their homes. However, when the lockdown was imposed, the testing had also been woefully inadequate, with a nationwide total of just 22,694 tests having been conducted upto that date. If we use the extrapolation technique of inferring case counts from death counts as we did in §5, then using the same 1 percent mortality rate and 20 day interval to death, we find almost 40,000 assumed cases on the day that the lockdown began. If we go by this figure, then the lockdown wasn’t really early, and possibly should have been enforced earlier still in trouble zones such as Mumbai. Certainly, if the figure of 40,000 cases is true, then one further week of normal life (with huge crowds in trains and railway stations) might have been disastrous. From the vantage point of today, alternate arrangements should definitely have been made much earlier for rehabilitation of the migrant workers. However these arrangements would have involved considerable complexity in the prevailing situation, and were certainly not as easy as one week’s delay in announcing lockdown.

Sweden, which has adopted a controlled herd immunity strategy, has been accused of playing with fire. It is also possible that the Swedish authorities are aware that they do not have the contact tracing capacity required for performing like City A and hence are attempting something like City D – a faster end of the epidemic than City B at the expense of a higher case count. To make a comprehensive analysis of their policy, it is crucial to know not only the last intricate detail of the epidemiological aspects but also the details of the economic considerations. That is almost impossible. On a different note however, we are seeing extremely recent reports [50], [51] stating that the virus has entered into old age homes and similar establishments, causing hundreds of deaths over there. Assuming that these reports are not overturned in the course of time, allowing the ingress of virus into high-risk areas is an indefensible action, whatever the overall epidemiological strategy.

Finally, extremely important public health factors such as the racial dependence of susceptibility and/or transmissibility have just started coming to the surface. Another complete grey area is the mutations which this new and vicious virus are undergoing and what effect they might have on the spreading dynamics. Some reports also reflect that the change in genetic composition due to mutation might be the reason behind huge differences in the crude infection rate between countries [52], [53]. In the absence of a clear picture about this, any public health measure is all the more likely to be a random guess with non-zero probabilities of both success and failure.

Not everything about corona is random or outside one’s control though. Amongst the European countries, we can see that Germany, Austria, Switzerland, Denmark, Norway and Finland have definitely managed the epidemic while their neighbours have not, which rules out some hidden luck factor. The same has happened in Kerala and Karnataka (also in India). This has been feasible only due to governmental awareness and hard work, and people’s cooperation. Similarly, there are some governments which have been clearly guilty of negligence or hubris in their management of the disease. It would also be noteworthy to observe and take lessons from the some of the new places like Alabama, Arkansas, etc which have been recently identified as potential hotspots of this pandemic. Finally, we note that our talk of parameter sensitivity does not imply a condonation of the actions of countries and supranational organizations whose suppression and manipulation of information has led to the pandemic’s acquiring its current, horrific proportions. However, between the obviously competent governments and the obviously negligent ones, there are several hundred intermediates and it is these whose smug, retroactive, backward-glancing criticism we ask that all of us refrain from. Coronavirus is not some kind of race but a public health disaster and we should adopt a unified approach to the fight against it.

## CONCLUSION

Here, we summarize the take-home messages of each Section of this Article.

- §1 – The global spread of corona has shown multiple anomalous features, the primary one being vastly disparate case trajectories in regions with similar strength of lockdowns.
- §2 – The above anomalies can be explained if the fraction of asymptomatic carriers is much higher than previously thought. In such a case, contact tracing becomes the most important factor.
- §3 – The model proposed in Ref. [2] is excellent when *R* < 1 but inapplicable when *R* > 1. It acts as the basis of the present, more advanced model.
- §4 – The retarded logistic equation (3) corrects the limitation mentioned above and we claim it to be the central ingredient of global Coronavirus dynamics.
- §5 – The credibility of the model (3) is enhanced through plausible fits of the case trajectories of hotspot regions.
- §6 – For different parameter values, (3) exhibits different classes of solutions. This Section together with §3 and §4 constitute the mathematical core of this Article.
- §7 – The reported case trajectories in various regions are also distorted by (*a*) limited testing capacity and (*b*) high false negative ratio of real time RT-PCR test.
- §8 – The spreading of the virus all over the world can be understood in terms of the different solution classes considered in §6.
- §9 – A city can reopen only if it is past the peak of cases. Reopening must be accompanied by robust contact tracing. The US CDC has laid down a set of reopening guidelines which are compatible with our model and its solutions.
- §10 – Efficiency of contact tracing comes at the expense of people’s privacy – balancing between the two is a delicate optimization problem.
- §11 – In some regions, restrictions such as masks and six-feet separation minima must be maintained for a very long time to come. The public health authorities can ensure compliance by resorting to behaviour-theoretic approaches.
- §12 – People of East Asian race have intrinsically lower susceptibility or transmissibility of this zoonotic virus.
- §13 – Small changes in parameter values in (3) can cause huge changes in the region’s case trajectories.

Six Wednesdays ago, we had concluded our previous Article [2] with a hope that COVID-19 may become history by the end of summer. In many regions of the world at least, the hope now appears fond. Contrary to our perceptions of having become a technocratic super-specie, humankind has proved remarkably powerless in the face of tribulation. It has taken a mere three months for the entire civilized world to retreat into a submissive state, desperately defending ourselves against this curse instead of combating it as an equal. Only That Which earth and seas obey can still take COVID-19 back in less time and with less destruction than what we have found, and we close this Article with a prayer that this may be the case.

## Data Availability

This Article uses data available from websites which are included in the References.

